# Industrialized complementary food for infants and young children: a systematic review protocol of their nutrient profile and impact on health outcomes

**DOI:** 10.1101/2022.09.19.22280098

**Authors:** Juliana Morais de Sousa, Priscila Gomes Oliveira, Elisa Maria Rodrigues da Silva, Nicolie Mattenhauer de Oliveira, Karla Danielly da Silva Ribeiro

**Author notes:** Corresponding author (KDSR). These authors contributed equally to this work (Juliana Morais de Sousa, Priscila Gomes Oliveira, Karla Danielly da Silva Ribeiro). These authors also contributed equally to this work (Elisa Maria Rodrigues da Silva, Nicolie Mattenhauer de Oliveira).

## Abstract

Complementary feeding plays an important role in the patterns of growth, development and formation of eating habits. Although the consumption of complementary foods (CF) with an inadequate Nutrient Profile (NP) is considered a risk factor for morbidities, there are still few studies that assess its repercussions on children’s diet and health. This review aims to identify the participation of industrialized CF consumption in children’s diet, the NP of these foods, and their impact on quality diet and on health of children under two years of age. This is a protocol study for systematic review registered in the International Prospective Register of Systematic Reviews (PROSPERO) CDR 42022321891, following recommendations of the Preferred Reporting Items for Systematic Review and Meta-analysis Protocols (PRISMA-P). We will seek out cross-sectional or cohort studies investigating the NP of industrialized CF recommended for children up to two years and/or that evaluated the association between the CF consumption and the children’s diet and health. The search for records will be conducted on PubMed/Medline, Scopus, Embase, Web of Science, and Scielo. Two independent reviewers will perform all steps of the systematic review. The methodological quality will be analyzed using the Newcastle–Ottawa scale (NOS) and Risk of Bias 2 (RoB 2). Results will be presented by means, medians, confidence interval (95%), standard deviation for the NP of foods, and to assess the health impact, comparisons of outcome measures, effect sizes (ORs and RRs) will be extracted. The high consumption of industrialized CF and the negative impact on children’s health, in addition to the gap in the literature of studies assessing the NP and consumption of these foods by children under 24 months, justify the importance of a review on this subject focusing on that age group.

## Introduction

The first two years of life constitute a critical window for promoting child health and developing healthy eating practices^1,2^. Complementary feeding describes a period in which there is a gradual reduction in the frequency and volume of breast milk or infant formula, along with the introduction of complementary foods (CF)^3^. The quantity, frequency, variety, and consistency of CF must be adequate to meet nutritional needs^4^.

CF inadequate qualitative profile, such as an excessive intake of calories, salt, sugars and fats, are a threat to health as they increase the risk of chronic diseases and micronutrient deficiency^5^. Complementary feeding plays an important role in the pattern of growth, development, in the configuration of eating preferences and behaviors that can lead to the development of overweight, obesity and other noncommunicable diseases (NCDs) in childhood and adulthood^3,6,7^.

Childhood obesity affects children’s physical, social and psychological well-being. It is considered a direct cause of childhood comorbidities, especially cardiometabolic diseases, and may constitute a risk factor for obesity in adulthood^8^. It is estimated that 6.7% of children under 5 years of age were overweight in the year 2020, caused in part by changes in food availability and quality combined with a reduction in physical activity levels^8^. Exposure to obesogenic environments and the aforementioned factors result in energy imbalance, favoring childhood overweight^9^. There is growing concern that the consumption of breastmilk substitutes and some commercial CF are harming infant feeding and contributing to overweight. The early introduction of CF has been associated with breastfeeding interruption and with lower consumption of fresh and/or minimally processed foods, leading to a reduction in dietary quality^1,2,10^.

Most foods marketed to infants are considered processed and ultra-processed (UPF), such as infant formulas and cereals, and have a NP of higher energy density, free sugars, total or saturated fats, and less fiber when compared to fresh foods^11,1,4^. More than 80% of children under two years of age have already been exposed to some type of industrialized food^12,13^.

Foods such as snacks, cakes, bread, breakfast cereals and sugary drinks are the most consumed^14,15,16^. The consumption of sugary drinks is considered high in the pediatric age group, with already consumed by 95% of children in a European cohort aged 9-12 months^6^ and more than half of children aged 12 to 23 months in low- and middle-income countries^17^.

It is noteworthy that the consumption of processed foods in childhood with excess of critical nutrients^18^ (PAHO) is related to different health and nutrition outcomes, such as inadequate eating practices, poorer diet quality, excess measures and indicators of adiposity, changes in lipid profile, respiratory diseases, dental caries and even toxicity related to the ingestion of plastics present in food packaging^16,19,20^.

With the objective of reducing the accelerated growth of obesity and preventing the emergence of NCDs throughout life the WHO instituted a set of interventions to modify the obesogenic environment, some being the regulation of the marketing of food and beverages targeted at children, food labeling policies, healthy eating promotion, and protection of breastfeeding^9^.

Despite the growing number of publications on the consumption of processed foods by the population, studies with children are limited in number and methodological quality ^19,21,22^, especially in the age group under 2 years, thus being necessary to assess the impact of consumption of unhealthy foods and beverages on nutritional status and child growth^23^. Given this gap in literature and the severity of the impact of processed food consumption on children’s health, our review aims to identify: a) the participation of industrialized complementary foods in the diet of children, b) the NP of these foods, and c) the impact of the consumption of these foods on the quality diet and on the health of children under two years of age.

## Materials and Methods

### Protocol and registration

This protocol is registered in the International Prospective Register of Systematic Reviews (PROSPERO) (CDR 42022321891) and follows the recommendations of the Preferred Reporting Items for Systematic Review and Meta-analysis Protocols (PRISMA-P)^24^. To ensure the quality of the protocol, the PRISMA-P checklist (S1 File) was used. During the review, changes may be made to the protocol, which will be duly justified and disclosed through PROSPERO.

### Review question

The research question was formulated based on the acronym PEO, which considers P, population; E, Exposure; O, result (Table 1), following the question: “What is the impact of the consumption of industrialized complementary foods on children’s diet and health?”

**Table 1:** Definition of the structured review question based on PEO.

| PEO | Description of the item in our study |
| --- | --- |
| P<br>(Population) | - Children aged 0-24 months OR<br>- Commercial complementary food (CF) for 0-24-month-old children. |
| E<br>(Exposure) | - Commercial complementary food |
| O<br>(Outcome) | - Nutritional composition of commercial CF,<br>- Impact of processed CF consumption on the quality diet,<br>- Impact of processed CF consumption on children's health. |

### Inclusion criteria

In the review, observational studies with cross-sectional design or cohort studies will be considered eligible. Studies which evaluate commercial CF recommended for 0-24-month-old children will be included, including those which only evaluate the NP of the food itself, as well as those that evaluate CF consumption by children and their impact on health.

For the definition of “industrialized complementary foods”, processed and ultra-processed foods will be considered. Processed foods are those composed of foods or part of processed foods, containing added salt, sugar, oils or fats, and ultra-processed food (UPF) are defined as formulations of ingredients derived from foods and additives, combined with substances that include colorings, flavorings, sweeteners and emulsifiers – food substances of no or rare culinary use according to the NOVA classification^25,26^. Studies must clearly state in their text that the analyzed food is considered an industrialized CF.

The NP will consider the following criteria: Macronutrient and micronutrient composition (g/mg/μg), Diet Quality Index (DQI), nutritional adequacy (% EAR – Estimated Average Requirement), breastfeeding prevalence (% Exclusive Breastfeeding – EB; % Breastfeeding – BF in 0-6-month-old infants; % Continued Breastfeeding in 0-6-month-old infants).

Food will be evaluated according to the groups: Biscuits/wafers; Cereals/porridge; Fruit and/or vegetable puree; Juice/Smoothie/Tea/Other Beverages (ready to drink); Meat- or fish-based ready-to-eat meal; powdered milkshake; Puree dessert (e.g., pudding, cream); Soup; Dairy compounds; Yogurt or yogurt-related product, and Others^4^. We will not consider breastmilk substitutes (infant formulas for 0-6-month-old children, follow-up formulas for 6-12-month-old children and growth milk, dairy compounds for 0-36-month-old children).

For health outcomes, the following will be considered: indicators of anthropometric nutritional status (z score of anthropometric indices), adiposity measures, biochemical analysis (mg/μg/dL), and disease indicators (presence or absence).

### Exclusion criteria

The following will be excluded: (1) reviews, conference abstracts, opinions, experimental studies, research protocols, case controls, clinical trials, case reports, comments, letters to editors, (2) studies which did not consider consumption and/or commercialization of industrialized CF for 0-24-month-old children, (3) studies which did not include a separate analysis of the target population (0-24-month-old children), and (4) studies which only considered infant formula/breast milk substitutes as industrialized CF.

### Search strategy

The search strategies used in the identification phase of the studies will be based on the definition of descriptors (MeSH) and their equivalents, related to industrialized CF consumed by and/or marketed to 0-24-month-old children (Table 2).

**Table 2:** Search equations for conducting a systematic review.

| PEO | Descriptors |
| --- | --- |
| P<br>(Population) | "Infant" [MESH] (Children OR Child) |
| E<br>(Exposure) | "Infant food" [MESH] (Infants foods OR infant food OR infant foods OR baby food OR complementary foods OR complementary food OR complementary feeding OR industrialized food OR processed food OR ultra-processed food) |
| O<br>(Outcome) | "Outcome health" [MESH] (Outcome Assessment OR Outcomes Assessment) OR "Eating" [MESH] (Food Intake OR Intake, Food OR Dietary Intake OR Ingestion) OR "Nutritive Value" [MESH] (Nutritive Values OR Nutritional Value OR Nutrition Value OR Nutrition Values OR Nutritional Quality OR Nutritive Quality OR Nutritional Food Quality OR Food Quality, Nutritional) |

The search for records will be conducted on the following databases: PubMed/ MEDLINE, SCOPUS, EMBASE, Web of Science and SciELO, based on eligibility criteria. The language of publication will be limited to English, Portuguese, French and Spanish. There will be no limitation on the year of publication.

Researchers will also verify the reference list of studies included in the review to add eligible studies not retrieved by the search strategy. A new search will be performed at the end of the review in order to retrieve the most recent studies eligible for inclusion in the review. Missing data were sought via emails to the corresponding authors or from secondary publications of the same study. Records will be download to Rayyan plataform, added to a standardized data collection form and duplicates were removed.

### Study selection and data extraction

All stages of the systematic review (search, eligibility criteria analysis, data extraction and study quality appraisal) will be performed by four independent reviewers (JMS and PGO, EMRS and NMO). Records selected according to the search strategy will be sent to the Rayyan QCRI application (www.rayyan.ai) for the initial screening of titles (phase 1) and abstracts (phase 2) respectively, according to the eligibility criteria. When there are differences between the judgments among the four reviewers, the conflicting decision will be resolved by a fifth reviewer with extensive scientific knowledge on the topic (KDSR). Subsequently, the two researchers will analyze the full text of the records filtered in the previous phases to select the studies according to the eligibility criteria.

The PRISMA-P flowchart will be used to present the studies that were selected according to the eligibility criteria (Fig 1).

**Figure 1:**
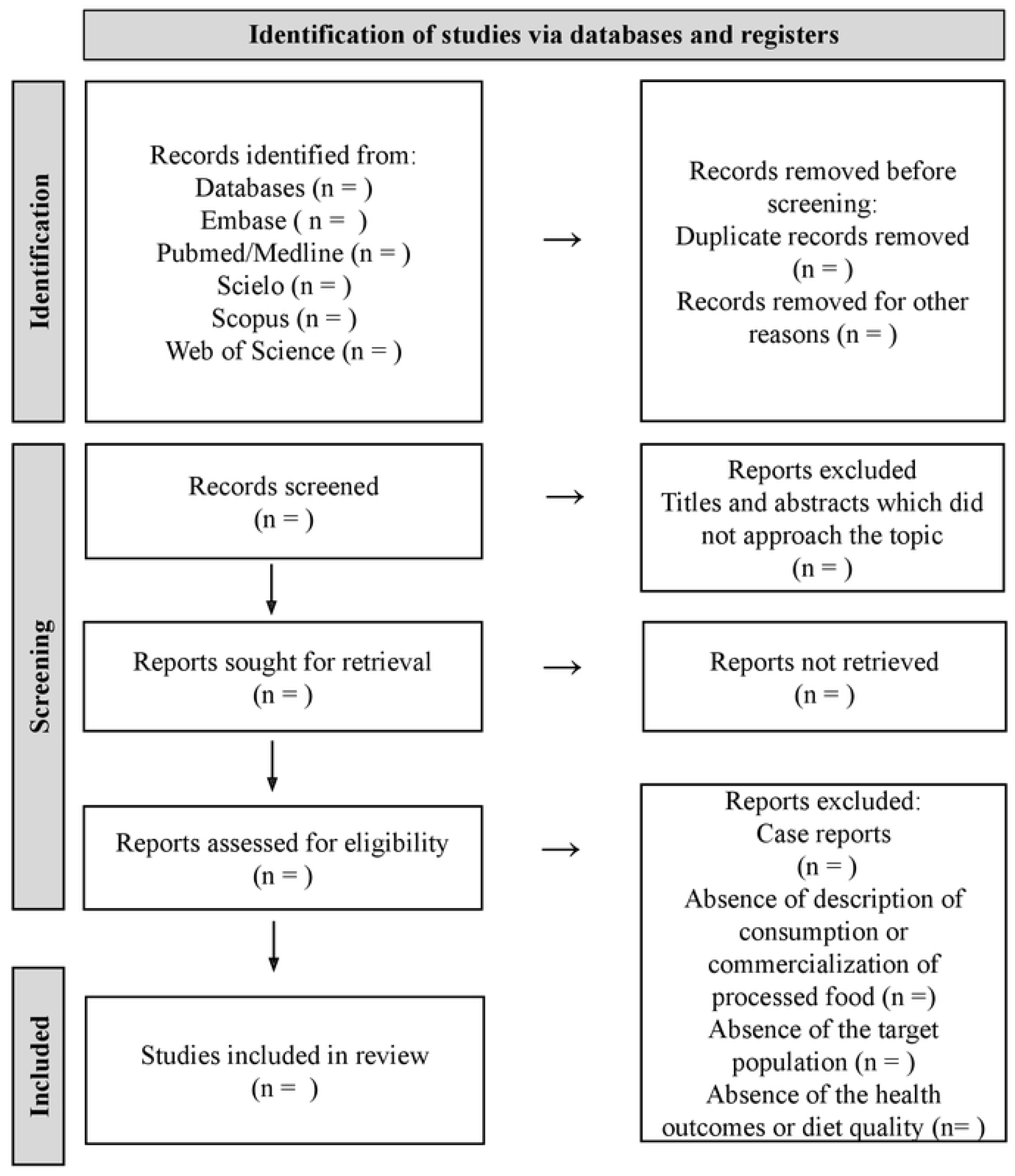
Flow diagram of the study selection based on PRISMA-P

### Data extraction

Data extraction will be done with a standardized form divided into two sessions:

A. NP of industrialized CF, according to food type: study country, type of study, number of foods (= n), target age group, food analysis instrument, nutritional composition by type of food (energy, macronutrients, sugars and sodium - kcal/ g/mg/).
B. Consumption of industrialized CF and impact on health: study site, type of study, number of study participants (= n), age group, exposure to processed foods (% of calories consumed or number of foods consumed per day), CF types, food survey types, health outcomes (body mass index for age (BMI/A), waist circumference, adiposity - z escore, cm, mm), diet quality, and main study findings.

### Risk of bias and quality assessment

The methodological quality of each included study will be independently assessed by two reviewers (JMS and PGO). Any discrepancies in decisions will be resolved by consultation with a third reviewer (KDSR). The Newcastle–Ottawa (NOS) scale will be used to assess the methodological quality of retrospective studies, applying the modified version (Risk of Bias Tool 2 – RoB 2) to assess cross-sectional studies included in the review. The tools assess biases from confounders, participant selection (sampling), missing data, and outcome measurement. Study authors will be contacted in the event of insufficient detail to confidently assess the risk of bias.

### Strategy for data synthesis

In the nutrient profile axis of CF, the means, medians, confidence interval (95%) and standard deviation of the nutritional composition values will be used, according to the type of food analyzed.

For the axis impact of consumption of commercial CF on health and diet quality, comparisons of outcome measures, effect sizes (ORs and RRs) with CIs of 95% will be extracted.

## Discussion

CF commercialization is growing and expressive. Foods with an inadequate nutritional quality are commercialized to infants and young children^1,4,27^. Commercially available CF are highly heterogeneous, varying substantially in energy and nutrient density^27^. The early introduction of processed foods and insufficient consumption of fresh foods can have negative impacts on children’s health^36,37^. There is heterogeneity in the profile of such foods, regarding processing technique and nutritional composition. Such information needs to be considered in epidemiological studies which assess the association of UPF consumption with adverse health outcomes in early childhood^38^.

Aiming at protecting breastfeeding, preventing obesity, chronic diseases and promoting healthy eating, the WHO has released guidelines to end the inappropriate promotion of food and beverages aimed at 0-36-month-old children. According to the guide, complementary feeding should be timely, start at 6 months of age, must be adequate and offer CF in the correct quantity, frequency, consistency and variability and must be hygienically and sanitarily safe. Food for infants and young children, other than breastmilk substitutes, may be promoted as long as they meet the quality and nutrient composition established in national dietary guidelines^1^.

In 2021, the WHO published new indicators of dietary practices^28^, highlighting two of them aimed at identifying inappropriate dietary practices: the consumption of foods considered unhealthy and the consumption of sweetened beverages among children aged 6 to 23 months. Early consumption of sugary drinks is associated with the risk of childhood obesity and tooth decay, and consumption of unhealthy foods can replace more nutritious foods and limit the intake of essential vitamins and minerals.

Some studies point to a greater availability in the market of CF for infants of the UPF type, such as infant formulas, dairy compounds, ready-to-eat meals and cereals, which have higher energy, carbohydrate and fat content, and lower fiber content than minimally processed foods^11^. But there is still a need to assess the profile of these foods according to their type.

UPF correspond to 42% of children’s energy intake in Brazil^31,32^, 48% in Canada^33^, 67.8% in England^34^ and 76% in the United Kingdom^35^.

From a consumption perspective, ultra-processed CF intake is associated with diminishing of the diet quality, being negatively correlated with fiber and protein, and positively with the intake of sugar, total fat and saturated fat^29^. Studies have showed that Brazilian children who were breastfed exclusively for less than 4 months had lower fruits and vegetables consumption and higher consumption of processed foods during food introduction, with almost half of the energy consumed (43%) coming from processed foods and UPF^30^.

Among the possible limitations of our systematic review, the heterogeneity of the included studies and the different foods and methods for measuring outcomes related to nutritional status and diet quality stand out. To minimize these biases, we will use strategies to standardize measures and indicators (data synthesis), profile presentation according to the type of food, inclusion only of studies that clearly stated that they are processed foods, and of those that include the % of their participation in the diet, presented by country, which will make possible for us to provide systematic information on CF.

The Brazilian dietary guidelines for children under 2 years old^2^ reinforces the importance of continued breastfeeding and the consumption of fresh or minimally processed foods during complementary feeding, however, what is observed in practice is an early introduction of processed foods. This excessive and early consumption can lead to negative effects on children’s health with repercussions in adult life^21^. Although there are already studies that address the impacts of the consumption of processed foods in childhood, there is a gap in the literature of studies that assess the NP of these foods intended for and/or consumed by 0-24-month-old children^19^, which justifies the relevance of a new review on this topic.

## Data Availability

No datasets were generated or analysed during the current study. All relevant data from this study will be made available upon study completion.

## Supporting information

**S1 Fig 1. Flow diagram of the study selection based on PRISMA-P** (PDF)

## Author contributions

### Conceptualization

Juliana Morais de Sousa, Priscila Gomes Oliveira, Karla Danielly da Silva Ribeiro

### Formal analysis and corresponding reviewer

Karla Danielly Silva Ribeiro

### Investigation

Juliana Morais de Sousa, Priscila Gomes Oliveira, Elisa Maria Rodrigues da Silva, Nicolie Mattenhauer de Oliveira, Karla Danielly Silva Ribeiro.

### Validation

Juliana Morais de Sousa, Priscila Gomes Oliveira, Karla Danielly Silva Ribeiro

### Visualization

Juliana Morais de Sousa, Priscila Gomes Oliveira, Elisa Maria Rodrigues da Silva, Nicolie Mattenhauer de Oliveira, Karla Danielly Silva Ribeiro

## References

1 World Health Organization. Guidance on ending the inappropriate promotion of foods for infants and young children: implementation manual. Geneva: WHO; 2017.

2 Brazil, Ministry of Health. Food Guide for Children Under 2 Years. Brazil : Ministry of Health ; 2019

3 D’Auria E et al. Complementary feeding: Pitfalls for health outcomes.International Journal of Environmental Research and Public Health. 2020; 17(21): 7931.

4 World Health Organization. Commercial foods for infants and young children in the WHO European Region: A study of the availability, composition and marketing of baby foods in four European countries. Geneva: WHO; 2019

5 Lutter CK, Grummer-Strawn L, Rogers L. Complementary feeding of infants and young children 6 to 23 months of aige.Nutr Rev. 2021; 79(8): 825–846.

6 Theurich MA et al. Commercial complementary food use amongst European infants and children: results from the EU Childhood Obesity Project. European Journal of Nutrition. 2020; 59(4): 1679–1692.

7 Rodríguez-Cano et al. Complementary feeding practices and their association with adiposity indicators at 12 months of age. Journal of Developmental Origins of Health And Disease. 2021; 12(5): 780–787.

8 Lobstein T, Brinsden H, Neveux M. World Obesity Atlas 2022. London: World Obesity Federation; 2022.

9 World Health Organization. Report of the commission on ending childhood obesity. Geneva: WHO; 2016.

10 World Health Organization. Global strategy for infant and young child feeding. Geneva: WHO; 2003.

11 Da Rocha KF et al. Commercial foods for infants under the age of 36 months: an assessment of the availability and nutrient profile of ultra-processed foods. Public Health Nutrition. 2021; 24(11): 3179–3186.

12 Giesta JM et al. Associated factors with early introduction of ultra-processed foods in feeding children under two years old. Science & Health Collective. 2019; 4: 2387–2397.

13 Cainelli EC et al. Ultra-processed foods consumption among children and associated socioeconomic and demographic factors. Einstein. 2021; 19:1–8.

14 Neri D et al. Consumption of ultra-processed foods and its association with added sugar content in the diets of US children, NHANES 2009-2014. Pediatric Obesity. 2019; 14(12): e12563.

15 Onita BM et al. Eating context and its association with ultra-processed food consumption by British children. Appetite. 2021; 157: 105007.

16 Neri D et al. Ultraprocessed food consumption and dietary nutrient profiles associated with obesity: A multicountry study of children and adolescents. Obesity Reviews. 2022; 23: e13387

17 Huffman SL et al. Babies, soft drinks and snacks: a concern in low- and middle-income countries?. Maternal and Child Nutrition. 2014; 10(4): 562–574.

18 World Health Organization. Pan American Health Organization Nutrient Profile Model. Washington, DC : PAHO, 2016.

19 Oliveira PG et al. Impacts of Consumption of Ultra-Processed Foods on the Maternal-Child Health: A Systematic Review. Frontiers in Nutrition. 2022; 9: 821657.

20 Marino M et al. A systematic review of worldwide consumption of ultra-processed foods: findings and criticisms. Nutrients. 2021; 13(8): 2778.

21 Louzada MCL et al. Impact of the consumption of ultra-processed foods on children, adolescents and adults’ health: scope review. Health Notebooks _ public. 2022; 37: e00323020

22 English LK et al. Types and amounts of complementary foods and beverages consumed and growth, size, and body composition: a systematic review. The American Journal of Clinical Nutrition. 2019; 109(Supplement_1): 956S–977S.

23 Rousham EK et al. Unhealthy Food and Beverage Consumption in Children and Risk of Overweight and Obesity: A Systematic Review and Meta-analysis. Advances in Nutrition. 2022; 00:1–28.

24 Moher D et al. Preferred reporting items for systematic review and meta-analysis protocols (PRISMA-P) 2015 statement. Systematic reviews. 2015; 4(1): 1–9.

25 Monteiro CA et al. A new classification of foods based on the extent and purpose of their processing. Health Notebooks _ public. 2010; 26: 2039–2049.

26 Monteiro CA et al. Ultra-processed foods: What they are and how to identify them. Public Health Nutrition. 2019; 22(5): 936–941.

27 Tzioumis E et al. Health effects of commercially-available complementary foods: a systematic review. Report to the World Health Organization; 2015.

28 World Health Organization. Indicators for assessing infant and young child feeding practices: definitions and measurement methods. Geneva; 2021.

29 Koiwai K et al. Consumption of ultra-processed foods decreases the quality of the overall diet of middle-aged Japanese adults. Public Health Nutrition. 2019; 22(16): 2999–3008.

30 Fonseca PCA et al. Association of exclusive breastfeeding duration with consumption of ultra-processed foods, fruit and vegetables in Brazilian children. European Journal of Nutrition. 2019; 58(7): 2887–2894.

31 Azeredo CM et al. Ultra-processed food consumption during childhood and asthma in adolescence: data from the 2004 Pelotas birth cohort study. Pediatric Allergy and Immunology. 2020; 31(1): 27–37.

32 Costa CS et al. Ultra-processed food consumption and its effects on anthropometric and glucose profile: a longitudinal study during childhood. Nutrition, Metabolism and Cardiovascular Diseases. 2019; 29(2): 177–184.

33 Moubarac JC et al. Consumption of ultra-processed foods predicts diet quality in Canada. Appetite. 2017; 108: 512–520.

34 Chang K et al. Association between childhood consumption of ultraprocessed food and adiposity trajectories in the avon longitudinal study of parents and children birth cohort. JAMA Pediatrics. 2021; 175(9): e211573–e211573.

35 Rauber F et al. Ultra-processed foods and excessive free sugar intake in the UK: a nationally representative cross-sectional study. BMJ Open. 2019; 9(10): e027546.

36 Karnopp EVN et al. Food consumption of children younger than 6 years according to the degree of food processing. Journal of Pediatrics. 2017; 93: 70–78.

37 Henriques P et al. Health and Food and Nutritional Security Policies: challenges in controlling childhood obesity. Science & Health Collective. 2018; 23: 4143–4152.

38 Lorenzoni G et al. What Is the Nutritional Composition of Ultra-Processed Food Marketed in Italy? Nutrients. 2021; 13(7): 2364.

